# Seven day continuous ambulatory electrocardiographic telemetric study with pocket electrocardiographic recording device for detecting hydroxychloroquin induced arrhythmias

**DOI:** 10.1101/2020.11.23.20232116

**Authors:** Rohit Walia, Nanda Prabhakaran, Ashwin Kodliwadmath, O Budha Charan Singh, Vikas Sabbarwal, Abhimanyu Nigam, Kartik Vijay B, Venkatesh Srinivasa Pai

## Abstract

**BACKGROUND & AIMS:** The use of hydroxycholoroquin for COVID 19 treatment and prophylaxis raised issues concerning its cardiac safety owing to possibility of QT prolongation and arrhythmias.^1^ There was no study on long term electrocardiographic telemetry monitoring of patients taking hydroxychloroquin and we planned a continuous electrocardiographic holter telemetry of these patients for a period of seven days.

**Methods:** Healthcare workers taking hydroxycholoroquin as pre exposure prophylaxis, patients taking hydroxychloroquin were monitored by holter electrocardiographic telemetry with continuous beat to beat analysis for seven days with capacity to report any arrhythmic event or significant QT prolongation instantly to medical faculty.

**Results:** 25 participants with mean age 42.4 ± 14.1 years, 40% females. 20% patients needed to stop HCQ. Four patients developed QT prolongation > 500 ms and needed to stop HCQ, one patient had accelerated idioventricular rhythm and stopped treatment. one had short episodes of atrial fibrillation. No malignant arrhythmia or ventricular arrhythmia or torsades were noted. No episode of significant conduction disturbance and arrhythmic death noted. Baseline mean QTc was 423.96 ± 32.18 ms, mean QTc corrected at 24 hours 438.93 ± 37.95, mean QTc 451.879 ± 37.99 at 48 hours, change in baseline mean QTc to max QTc was 30.74 ± 21.75 ms at 48 hours. All those develop QTc prolongation > 500 ms were greater than 50 years of age.

**Conclusion:** Ambulatory telemetry ECG monitoring seems to detect early QT prolongation and stopping drug timely prevented malignant arrhythmias. HCQ seems to have less risk of QT prolongation in young healthy individuals.

## Introduction

The use Hydroxycholoroquin (HCQ) for COVID 19 treatment and prophylaxis raised issues concerning its cardiac safety owing to possibility of QT prolongation and arrhythmias.^1,2^ Hydroxychloroquine, have been in use for treatment of malarias, lupus, and rheumatoid arthritis. Quinidine a member of group to which HCQ belongs is one of most torsade points (TDP) causing drug.^3^ There are few reports of arrhythmic potential of HCQ have been reported but exact frequency of events are underreported ^3^ and the exact incidence of drug induced TdP with HCQ is unknown. Inhibition of iKr and resultant QT prolongation associated with HCQ which can induce TDP and cause sudden cardiac death.^3,4^ There is lack of reliable data on this subject and risk owing to widespread unsupervised use in present day COVID pandemic is high. There was no study on long term electrocardiographic monitoring of patients taking HCQ and we planned a continuous electrocardiographic telemetry of these patients for a period of seven days.

## AIMS & OBJECTIVES

1. To assess QT prolongation to dangerous levels in patients on hydroxyclroroquin
2. To detect cardiac arrhythmias early and take preventive measures to prevent sudden cardiac death

### Methods

We did a prospective observational cohort study at tertiary care hospital. Healthy healthcare workers taking pre exposure COVID 19 HCQ prophylaxis, patients taking HCQ for any indication with written informed consent were included. The decision to take HCQ as prophylaxis and treatment was based on hospital COVID policy and patient preference. Those having a corrected QT > 480 ms, known congenital long QT, people having co morbidities and contra indicated to HCQ like retinopathy, known hypersensitivity to chloroquine, cardiomyopathy, prolonged QTc, cardiac arrhythmias, history of psoriasis, porphyria cutanea tarda, epilepsy, myasthenia gravis, myopathy of any cause, serious hepatic or renal disease, known glucose-6-phosphate dehydrogenase deficiency, current use of medication with known serious hepatotoxic effects or known interaction with chloroquine, severe depression, electrolyte imbalance, anti arrhythmic drugs, cardiac devices like pacemaker, defibrillators. The Indian Council of Medical Research, under the Ministry of Health and Family Welfare, has recommended chemoprophylaxis with hydroxychloroquine (400 mg twice on day 1, then 400 mg once a week thereafter) for asymptomatic healthcare workers treating patients with suspected or confirmed COVID-19, and for asymptomatic household contacts of confirmed cases. For treatment hydroxychloroquin 400 twice a day for first day followed by 400 mg once a day for 10 days was used.^5^

Pocket ECG (Medicalalgorithmics) constantly captures and classifies every heart-beat, records the onset and offset of every arrhythmia, detects all ventricular and supra ventricular arrhythmias, making for more comprehensive and complete data of home heart rhythm monitoring.

Monitoring methods that capture a full-disclosure ECG signal for every heart beat provide the most complete picture of a patient’s arrhythmia activity. By providing cardiologists online real-time access to diagnostic findings, these devices allow them to end a study once an arrhythmia is detected or extend it when further monitoring is required. Devices that also monitor physical activity enable cardiologists to differentiate between heart rate changes caused by physical activity and those caused by an arrhythmia.All clinical investigations were conducted according to the principles expressed in the Declaration of Helsinki (2001). The study protocol was approved by the Ethics Committee on Human Research of the institute and registered with clinical trails registry of India. All patients provided written informed consent before inclusion in the study. The study was registered with clinical trails registry of India.

#### Variables

Primary End Points were to analyse incidence of QT prolongation by 25% of baseline and number of patients with QT prolongation > 500 ms and with ventricular arrhythmias. Secondary Endpoints were to study drug related adverse effects. Continous electrocardiographic telemetry electrocardiography was used and physicians alerted immediately when dangerous levels of QT prolongation or arrhythmia events were noted.

#### Statistical methods

Continuous variables were reported as the mean ± SD and categorical variables were reported as numerical values and percentages. Paired t test was used to compare difference in QT c from baseline to maximum values.

### Results

#### Participants

Out of 200 healthcare workers only 21 gave informed consent to participate in study and were included and out of 100 patients admitted over a period of June to 1 October four patients gave informed consent and were included. The decision to take hydroxychloroquin as treatment or prophylaxis was based on hospital policy and various ongoing experimental treatments at our hospital by infectious disease department and once the patient has been decided to take hyroxychloroquin we asked them to participate in our study and took the willing patients. 25 people were recruited, 20 healthcare worker and 1 patient for pre exposure prophylaxis and 4 patients with COVID who were treated with HCQ.

Mean age was 42.4 ± 14.1 years, 40% (10)were female, 8%(2) had hypertension, 4 %(1) dyslipidemia and no case of diabetes or coronary artery disease (Table 1). Baseline mean QTc was 423.96 ± 32.18 ms, mean QTc corrected at 24 hours 438.93 ±37.95 (p value : 1.5), mean QTc 451.879 ± 37.99 at 48 hours (p value : 2.8), change from baseline mean QTc to maximum QTc was 30.74 ± 21.75 ms at 48 hours (Table 2). HCQ needed to be stopped in four patients because of QT prolongation. All significant QTc prolongation were seen in first 48 hours of start of therapy and subsided in next 3 days. When significant QTc was noted subjects were admitted and monitored till correction of QTc to normal values and removing and correcting other causes of QTc if present. No episode of TDP or malignant ventricular arrhythmia was recorded in any patient. One medical student had one episode of accelerated idioventricular rhythm and one person had short run of AF which seems to be not related to drug. Supra ventricular arrhythmias were not noted and few atrial premature complex (APC) and ventricular premature complex (VPC) within normal range were noted in study participants (Table 3).

**Table 1 :** Baseline Characteristics.

| **Age (years) (mean ± SD)** | 42.4 ± 14.1 |
| --- | --- |
| **Females** | 40%(10) |
| **Hypertension** | 8%(2) |
| **Diabetes** | Nil |
| **HCQ alone** | 24 |
| **HCQ + Azithromycin** | 4%(1) |

**Table 2 :** QTc Prolongation Trend.

| | QTc (Mean ± SD ) milli seconds |
| --- | --- |
| **Baseline** | 423.96 ± 32.18 |
| **QT c 24 hours** | 438.93 ±37.95 |
| **QT c 48 hours** | 451.879 ±37.99 |
| **Change QT c** | 30.74 ±21.75 ms |

Abbreviation - QTc: Corrected QT interval , SD : Standard deviation

**Table 3 :**
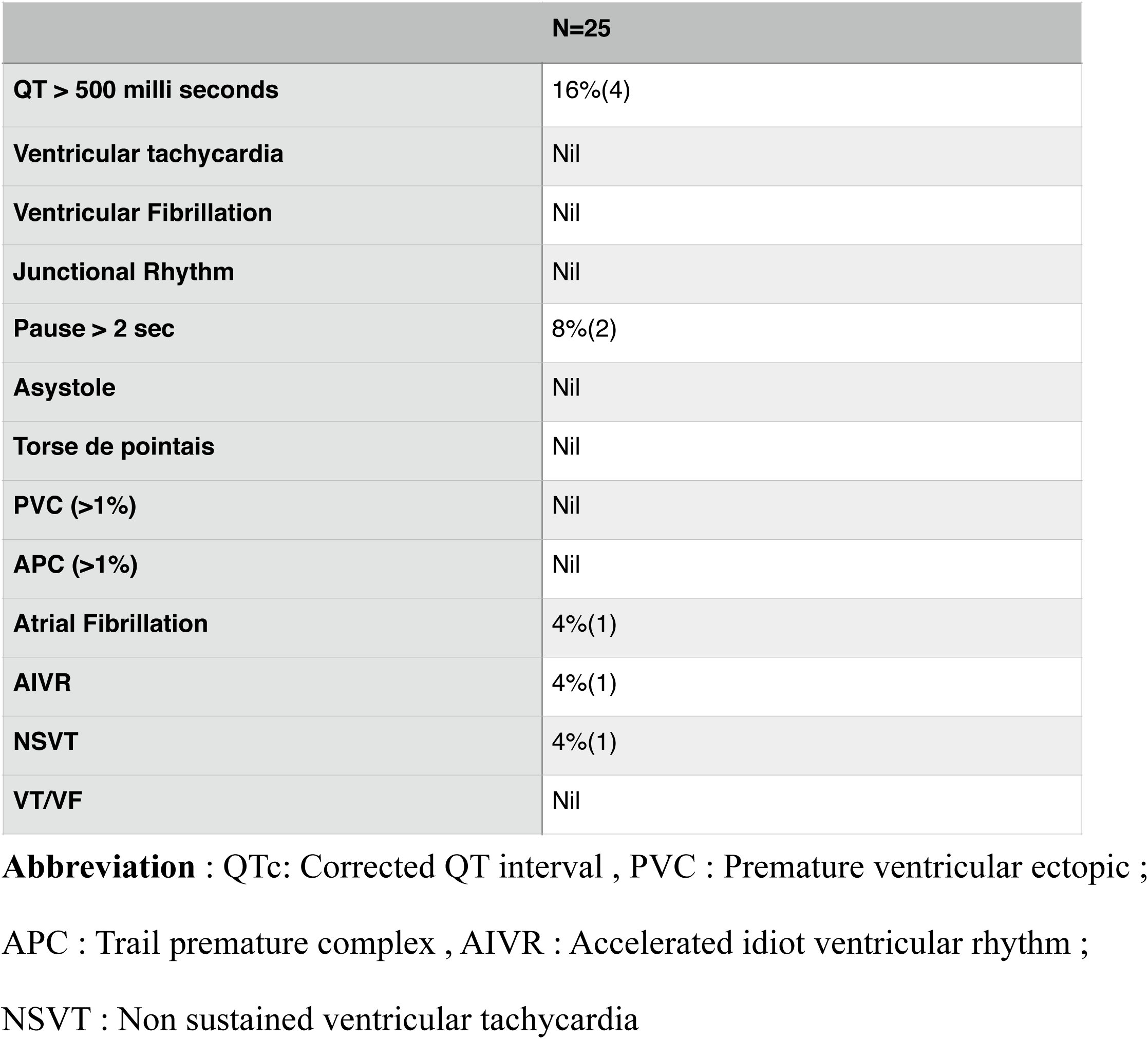
Arrhythmias and alerts.

All subjects developing QTc prolongation were greater than 50 years of age as compared to none in younger age group. Two were hypertensive. Most of prolongation of QTc occurred within 48 - 72 hours and reverted back within next 48 to 72 hours on stopping medication.

### Discussion

Indian Council of Medical Research (ICMR) Recommendations for electrocardiographic (ECG) monitoring for healthcare workers on HCQ prophylaxis is that an ECG (with estimation of QT interval) may be done before prescribing HCQ prophylaxis, in case any new cardiovascular symptoms occurs (e.g., palpitations, chest pain syncope) during the course of prophylaxis, An ECG (with estimation of QT interval) may be done in those who are already on HCQ prophylaxis before continuing it beyond 8 weeks, One ECG should be done anytime during the course of prophylaxis.^5^ There can be serious adverse events in following this protocol.

Cardiac Telemetry can detect early QT prolongation and can help in timely stopping the drug and take preventive measures. In elderly, hypertensives and those on diuretics or concomitant use of QT prolonging drugs may have more risk of QT prolongation and should be monitored closely. ^6^ HCQ did not produced any episode of TDP, significant supra ventricular or ventricular arrhythmias if QT prolongation is detected early and further medication is stopped.

Evidence from past studies has shown hydroxychloroquin and chloroquine, decreased excitability and conductivity of atrial and ventricular myocardium, though to lesser extent than quinine or quinidine.^7^ In a study of 28 patients taking 250 mg daily of chloroquine found QTc interval lengthened from 363–388 milliseconds to 372–392 milliseconds.^7^ But due to higher dose used in prophylaxis and treatment we observed more QTc prolongation. A study of 72 subjects with and without structural heart disease given acute chloroquine and hydroxychloroquine therapy for various types of atrial and ventricular arrhythmias observed one sudden death.^8^ hydroxychloroquine/chloroquine and torsades de pointes and ventricular tachycardia (VT) have been reported in this setting.^9,10^ A trial administering high doses of chloroquine (600 mg twice daily) in conjunction with azithromycin in suspected cases of severe COVID-19 pneumonia was stopped due to excessive QTc prolongation and association with increased mortality.^10^ Therefore, precautionary measures are necessary to mitigate the risk of QTc prolongation.^11^ It is important that in all previous studies on ECG there can be sudden deaths and continuous mobile telemetry with alerts and beat to beat report transmitted to physician through satellite based transmission is important to prevent sudden cardiac death. Monitoring methods that capture a full-disclosure ECG signal for every heart beat provide the most complete picture of a patient’s arrhythmia activity seems to be more protective than just ECG done at baseline and once a two or three day schedule.

## INTERMITTENT ECG VS TELEMETRY

Generally, drugs with potential QT prolongation should be avoided in patients with QTc greater than 500 milliseconds (or 550 milliseconds with intraventricular conduction delay) especially in the setting of moderate to severe structural heart disease and patients with high-grade atrioventricular block without a pacemaker and/or implantable cardioverter defibrillator in place.^12,13^ In our study we find the incidence of QTc prolongation to > 500 ms only in people greater than 50 years of age. The incidence was more in hypertensives and on concomitant QTc prolonging drugs. A 54year hypertensive female being treated with tablet Olmesartan 20mg once daily, tablet amlodipine 5mg once daily and tablet hydrochlorothiazide (HTZ) 12.5mg once daily. Baseline QTc was 431 ms calculated by Bazzet’s formula. Her baseline serum potassium was 4.1meq/l. After receiving 400 mg HCQ, the QTc on telemetry was 490 ms (Figure 1A) and after receiving 2 doses of HCQ 400 mg along with antihypertensive medications QTc rose to 530 ms (Figure 1B). Blood investigations showed a serum potassium of 3.48meq/L and serum magnesium of 2.08meq/L. She was admitted for continuous cardiac rhythm monitoring. The Tisdale score calculated for the patient was 8, indicative of moderate risk of drug induced QT prolongation. HCQ and hydrochlorothiazide were stopped as both have a tendency for QTc prolongation. Potassium replacement was begun with syrup potassium chloride 4tsp 6^th^ hourly and injection magnesium sulfate 0.5g intravenous (IV) 6^th^ hourly. Continuous cardiac monitoring could detect occasional ventricular premature complexes(VPC) but no polymorphic ventricular tachycardia(PVT) was detected and the patient remained asymptomatic. Twenty-four hours after the 2^nd^ dose of HCQ, the QTc was still 512 ms (Figure 1C). Regular monitoring of serum potassium and serum magnesium was done which showed an improving trend. After 57 hours of receiving the 2^nd^ HCQ dose, the serum potassium was 4.5 meq/L, serum magnesium was 2.56meq/L and telemetry showed a QTc of 453 ms (Figure 1D). The patient was discharged on oral magnesium salts and a high potassium diet, with withdrawal of hydroxychloroquine and hydrochlorothiazide from her medical regimen. Hydrochlorothiazide, a thiazide diuretic commonly used to treat hypertension, is known to cause side effects like hypokalaemia, hypomagnesimia and QTc prolongation, which maybe influenced by single nucleotide polymorphisms.^14^ The Tisdale score is used to predict the risk of QTc prolongation in patients being treated with QTc prolonging drugs. A Tisdale score of ≤ 6 predicts low risk, 7-10 medium risk, and ≥ 11 high risk of drug-associated QT prolongation.^2^

**Figure 1A.**
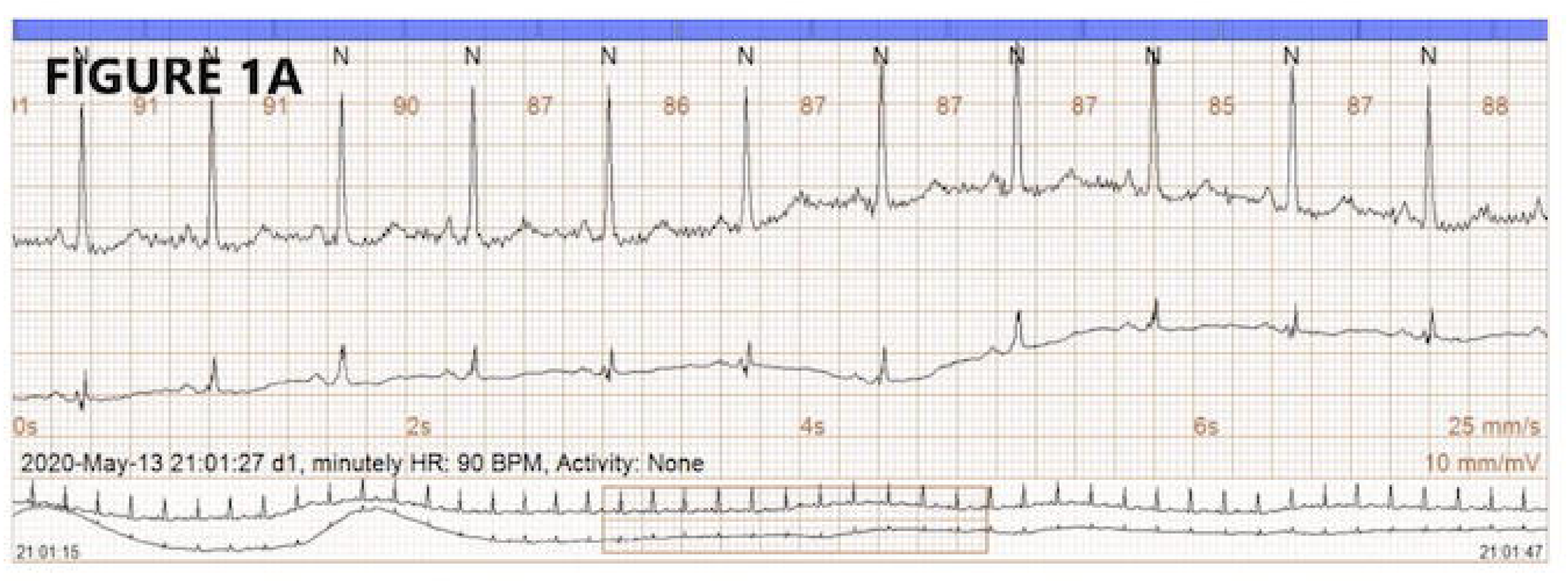
Telemetry ECG strip taken after receiving one dose of HCQ 400mg showing heart rate of 90bpm, QT interval of 400 ms and QTc interval by Bazzet’s formula of 490 ms.

**Figure 1B.**
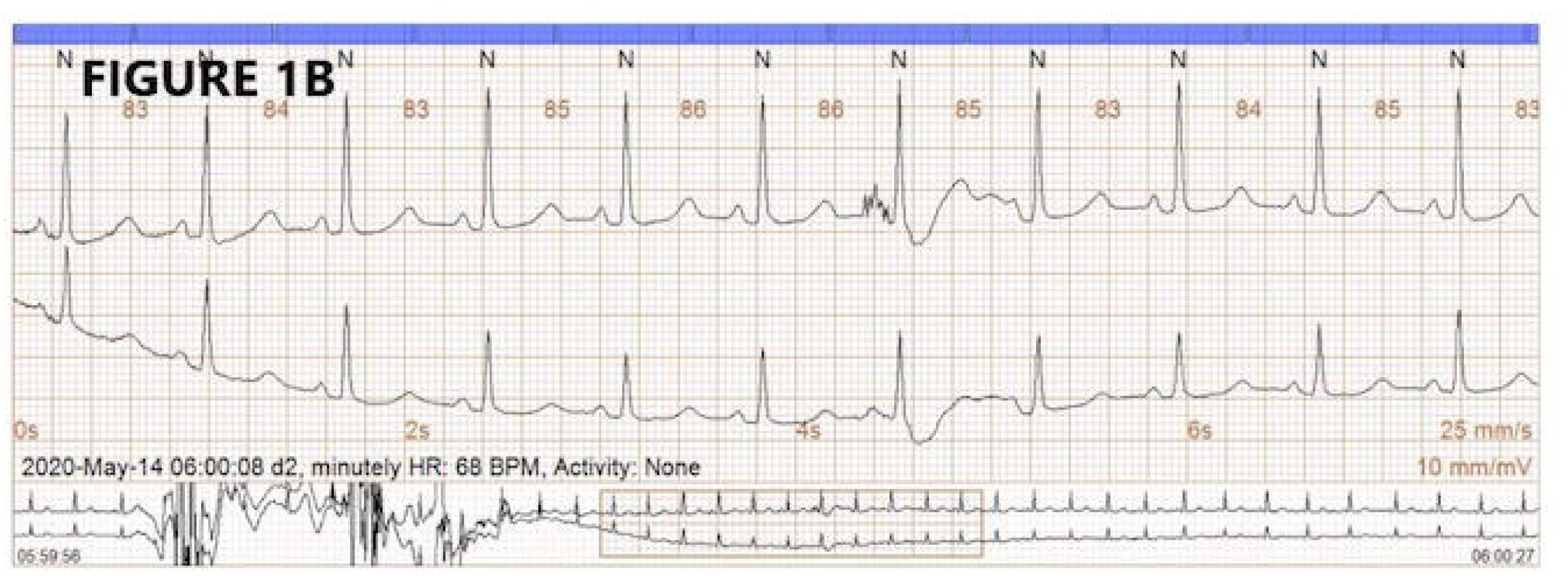
Telemetry ECG strip after taking 2 doses of HCQ 400 mg and 1 dose of HTZ 12.5 mg showing heart rate of 84bpm, QT interval of 448 ms and QTc interval by Bazzet’s formula of 530 ms.

**Figure 1C.**
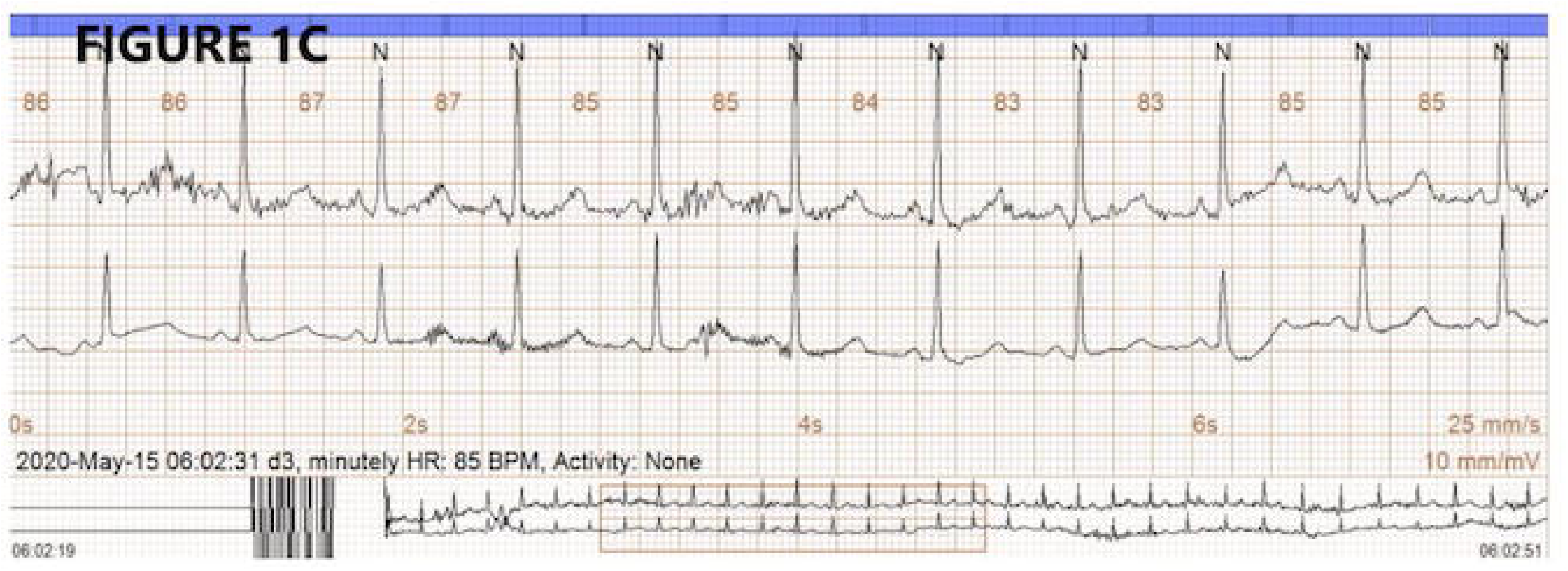
Telemetry ECG strip 24 hours after 2^nd^ dose of HCQ showing heart rate of 85bpm, QT interval of 430 ms and QTc interval by Bazzet’s formula of 512 ms.

**Figure 1D.**
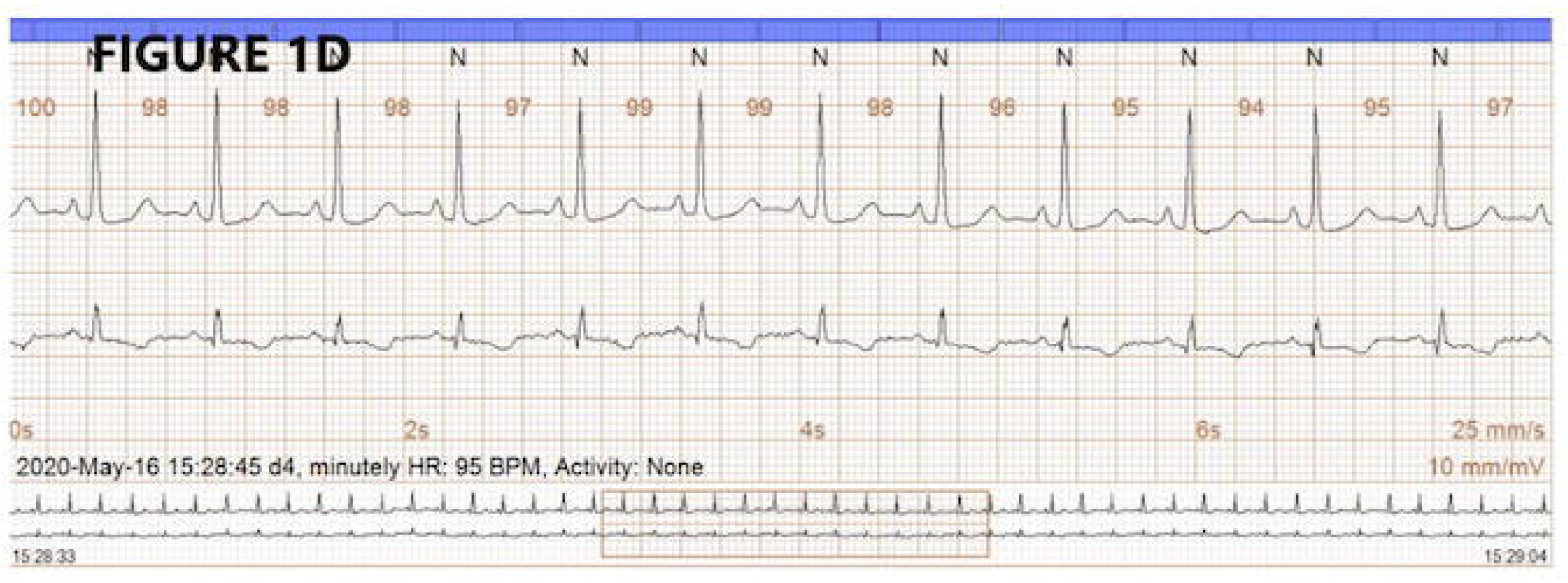
Telemetry ECG strip after 57 hours of 2^nd^ dose of HCQ showing heart rate of 95bpm, QT interval of 360 ms and QTc interval by Bazzet’s formula of 453 ms.

For patients on treatment with HCQ, it is recommended that the QTc should be monitored closely by an electrophysiologist

a. If QTc increases by >60 msec or absolute QTc > 500msec (or >530-550 msec if QRS >120 msec), reduce dose of hydroxychloroquine and repeat ECG daily.
b. If QTc remains increased >60 msec and/or absolute QTc >500 msec (or >530-550 msec if QRS >120 msec), reevaluate the risk/benefit of ongoing therapy, consider consultation with an electrophysiologist, and consider discontinuation of hydroxychloroquine.^2^

In contrast to intermittent ECG being used for monitoring and during treatment we found in our study that beat to beat telemetry based ambulatory monitoring is better.

### Repeated egg : more risk of transmission of infection

Though the role of mobile cardiac telemetry is controversial and frequent 12 lead ECG is recommended, it may not be possible to do frequent 12 lead ECG in all cases and thus mobile cardiac telemetry can lead to early detection and treatment. Intermittent ECGs miss timely detection of critical QTc prolongation and may result in malignant arrhythmias. Also frequent repeated ECGs are inconvenient to patients and not possible in ambulatory and remote health care workers monitoring. Frequent ECGs in infectious patients also carry a risk to infecting nursing staff and technicians.

## DURATION OF TELEMETRY AND RISK

All subjects developing QTc prolongation were greater than 50 years of age as compared to none in younger age group. Two were hypertensive. Most of prolongation of QTc occurred within 48 - 72 hours and reverted back within next 48 to 72 hours on stopping medication. So, this period should be at least monitored carefully ideally be telemetry.

### COVID 19 and Telemetry

In present COVID 19 pandemic and use of different drugs like resmedevir, faxiparin, lopinavir and ritonavir, azithromycin, etc which have potential to prolong QTc but not enough research data mobile based ambulatory telemetry seems to be ideal in detecting dangerous QTc prolongation and taking timely measures to prevent sudden cardiac death.

## CONCLUSION

Ambulatory continuous beat to beat telemetry seems to be advantageous in detecting timely and early changes in QTc interval to dangerous levels and prevent sudden death. This can be extremely valuable in present COVID 19 pandemic and use of numerous experimental drugs which have potential to prolong QTc and cause malignant arrhythmias.

### Limitations

The single-center, non-randomized study design, and a healthy population from a cardiac standpoint, are other limitations.

## Data Availability

Yes, all data is available with corresponding author and can be submitted as required.

## Disclosures

No conflict of interest to disclose

The study was approved by institutional ethics committee via letter number - AIIMS / IEC/20/255, dated 09/05/2020 and Clinical Trail Registry of India via letter number CTRI/2020/05/025216 [Registered on: 16/05/2020]

## Funding

Nil

## Notes

### Competing Interest Statement

The authors have declared no competing interest.

### Clinical Trial

Clinical Trail Registry of India, CTRI/2020/05/025216 [Registered on: 16/05/2020]

### Funding Statement

Non Funded study

### Author Declarations

All India Institute of Medical Science, Rishikesh

## REFERENCES

1. National Taskforce for COVID-19. Advisory on the use of hydroxy-chloro-quine as prophylaxis for SARS-CoV-2 infection. 2020. https://www.mo-hfw.gov.in/pdf/AdvisoryontheuseofHydroxychloroquinasprophylaxisforSARSCoV2infection.pdf (accessed March 23, 2020).

2. Mercuro NJ, Yen CF, Shim DJ, et al. Risk of QT Interval Prolongation Associated With Use of Hydroxychloroquine With or Without Concomitant Azithromycin Among Hospitalized Patients Testing Positive for Coronavirus Disease 2019 (COVID-19). JAMA Cardiol. 2020 May 1:E1–5.

3. Dapro B. Spectrum of drugs prolonging QT interval and the incidence of torsades de pointes. Eur Heart J 2001;3(suppl K):K70–80.

4. Yap YG, Camm AJ. Drug induced QT prolongation and torsades de pointes. Heart. 2003 Nov;89(11):1363–72.

5. https://www.mohfw.gov.in/pdfAdvisoryontheuseofHydroxychloro-quinasprophylaxisforSARSCoV2infection.pdf

6. Simpson TF, Kovacs RJ, Stecker EC. Ventricular Arrhythmia Risk Due to Hydroxychloroquine-Azithromycin Treatment For COVID-19. Am Coll Cardiol Magazine. 2020 Mar 29;00:1–9.

7. Naksuk N, Lazar S, Peeraphatdit TB. Cardiac safety of off-label COVID-19 drug therapy: a review and proposed monitoring protocol. Eur Heart J Acute Cardiovasc Care. 2020 Apr;9(3):215–221.

8. Wozniacka A, Cygankiewicz I, Chudzik M, et al. The cardiac safety of chloroquine phosphate treatment in patients with systemic lupus erythematosus: the influence on arrhythmia, heart rate variability and repolarization parameters. Lupus 2006; 15: 521–525.

9. Burrell ZL, Martinez AC. Chloroquine and hydroxychloroquine in the treatment of cardiac arrhythmias. N Engl J Med 1958; 258: 798–800.

10. Borba MGS, Val FFA, Sampaio VS, et al. Effect of high vs. low doses of chloroquine diphosphate as adjunctive therapy for patients hospitalized with severe acute respiratory syndrome coronavirus 2 (SARS-CoV-2) infection: a randomized clinical trial. JAMA Netw Open. 2020;3(4):e208857.

11. Giudicessi JR, Noseworthy PA, Friedman PA, Ackerman MJ. Urgent guidance for navigating and circumventing the QTc-prolonging and torsadogenic potential of possible pharmacotherapies for coronavirus disease 19 (COVID-19). Mayo Clin Proc. 2020;95(6):1213–1221.

12. Drew BJ, Ackerman MJ, Funk M, et al. Prevention of torsade de pointes in hospital settings. Circulation 2010; 121: 1047–1060.

13. Torp-Pedersen C, Møller M, Bloch-Thomsen PE, et al. Dofetilide in patients with congestive heart failure and left ventricular dysfunction. N Engl J Med 1999; 341: 857–865.

14. Seyerle AA, Sitlani CM, Noordam R, et al. Pharmacogenomics Study of Thiazide Diuretics and QT Interval in Multi-Ethnic Populations: The Cohorts for Heart and Aging Research in Genomic Epidemiology (CHARGE). Pharmacogenomics J. 2018 April; 18(2): 215–226.

